# Return of Results in a Global Sample of Psychiatric Genetics Researchers: Practices, Attitudes, and Knowledge

**DOI:** 10.1101/2020.06.08.20125641

**Authors:** Gabriel Lázaro-Muñoz, Laura Torgerson, Stacey Pereira

**Author notes:** Corresponding author: Gabriel Lázaro-Muñoz, PhD, JD, One Baylor Plaza, Suite 310D, Houston, TX 77346.

## Abstract

**Purpose:** Patient-participants in psychiatric genetics research may be at an increased risk for negative psychosocial impacts related to the return of genetic research results. Examining psychiatric genetics researchers’ return of results practices and perspectives can aid the development of empirically-informed and ethically-sound guidelines.

**Methods:** A survey of 407 psychiatric genetics researchers from 39 countries was conducted to examine current return of results practices, attitudes, and knowledge.

**Results:** Most respondents (61%) reported that their studies generated medically relevant genomic findings. Although 24% have returned results to individual participants, 52% of those involved in decisions about return of results plan to return or continue to return results. Respondents supported offering medically actionable results related to psychiatric disorders (82%), and the majority agreed non-medically actionable risks for Huntington’s (71%) and Alzheimer’s disease (64%) should be offered. About half (49%) of respondents supported offering reliable polygenic risk scores for psychiatric conditions. Despite plans to return, only 14% of researchers agreed there are adequate guidelines for returning results, and 59% rated their knowledge about how to manage the process for returning results as poor.

**Conclusion:** Psychiatric genetics researchers support returning a wide range of results to patient-participants, but they lack adequate knowledge and guidelines.

## INTRODUCTION

Psychiatric genetics has seen significant growth over the past decade.^1,2^ Much of this growth has been due to the expansion of large-scale genome-wide association studies (GWAS) for psychiatric disorders.^1,2,3^ During this time, psychiatry researchers have also started using more comprehensive single-nucleotide polymorphism (SNP) arrays, such as Illumina’s Infinium Global Screening Array, which allow psychiatric GWAS to generate medically relevant information related to both psychiatric and non-psychiatric conditions.^4,5^ The decreasing cost of genome-scale sequencing has also allowed an expansion of sequencing research in this field.^6,7,8^ The capacity to generate a quickly increasing number of medically relevant genomic findings in psychiatry research raises significant challenges. There is an emerging consensus that some medically relevant genomic findings should be offered to participants.^9,10^ Researchers in different fields, however, have struggled with how to responsibly manage medically relevant findings generated in their studies.^11,12,13^

A National Academies of Sciences, Engineering, and Medicine (NASEM) report and others have argued that determinations about returning results should be context-dependent.^14,15,16^ There has been little research on how to manage return of results in psychiatric genetics research, and even less on researchers’ experiences and perspectives.^17^ Studies have shown that stakeholders, including patients, caregivers, and clinicians, believe at least some results from psychiatric genetics studies should be offered to individual participants.^18,19,20^ Understanding researchers’ return of results practices, attitudes toward what, if any, findings should be offered, and perspectives on when and how to return results are especially important because of researchers’ roles in designing and allocating funds within studies, as well as their ultimate authority in determining how research findings are managed.

Return of individual results to participants in psychiatry research may accentuate certain challenges. Common concerns about return of research results are that participants may misunderstand the findings or that returning certain results could have a negative emotional impact.^13,21,22^ Given the focus of psychiatric genetics research, participants in these studies are more likely than in many other areas of genetics research to have mental health conditions that could affect their understanding of findings and potentially increase the likelihood that findings could negatively impact their emotional wellbeing. The impact of returning results to participants with psychiatric conditions, however, has not yet been thoroughly assessed, and there is little data available to guide the management of return of results to patient-participants in psychiatry research.^23^ At this point, it is also unclear what psychiatric genetics researchers’ current practices regarding return of results are, what, if anything, researchers’ believe should be offered to participants, and what their knowledge about return of results is. Thus, we surveyed a global sample of psychiatric genetics researchers to examine critical aspects about returning results to patient-participants. The results of this study can aid the development of guidelines to maximize the benefit and minimize the potential harms of returning genomic results to participants in psychiatry research.

## MATERIALS AND METHODS

### Participant Sampling

A web-based survey was administered using Qualtrics between July 2019 and December 2019 to members of the International Society of Psychiatric Genetics (ISPG), the largest international society dedicated to psychiatric genetics, and in person during ISPG’s World Congress of Psychiatric Genetics (WCPG) in October of 2019. Researchers were invited to participate via email or in person and were provided with personalized links. The Institutional Review Board (IRB) at Baylor College of Medicine approved the study. For those invited via email, reminders were sent at 2-3 week intervals up to 3 times. To incentivize participation, respondents could enter for a chance to win one of six $200 gift cards.

### Survey Measures

The survey was developed based on a review of relevant literature and themes that emerged during a prior study in which we interviewed 39 psychiatric genetics researchers from 17 countries to examine their perspectives on returning genomic research results.^10,17^ The survey instrument was a web-based questionnaire that took approximately 15-20 minutes to complete. We investigated the following domains: 1) roles and practices as a psychiatric genetics researcher; 2) knowledge about return of results; 3) attitudes; 4) challenges; 5) motivations; 6) ideal process for return of results to participants; and 7) demographics. We defined “medically actionable” in the survey as a result that indicates risk for a health condition for which there is some medical intervention available that can help decrease the risk of illness or help manage symptoms.^24^

Roles and practices were investigated in a series of multiple choice and yes-no questions to learn the researcher’s role, type of genetic testing used, participant populations studied, generation of medically relevant findings, and types of results returned, if any. Subjective knowledge questions asked respondents to rate their knowledge about various aspects of return of results on a 4-point scale from very poor to very good. Questions about attitudes, barriers, and motivations were explored using five-point agreement Likert scales (“strongly disagree” to “strongly agree,” with “neither agree nor disagree” as the midpoint). Two cognitive interviews^25^ with psychiatric genetics researchers were conducted to assess the web-based survey question relevance, readability, face validity, comprehension, and survey length. Minor changes were made to survey questions and answer choice format. The web-based survey was then tested by 10 individuals and piloted with 5 psychiatric genetics researchers. No changes were necessary based on the pilot.

### Data Analysis

Response frequencies are reported for each item. Differences in sample sizes reflect missing responses by respondents. When five-point or four-point Likert scale items were analyzed the two responses at each extreme (e.g., “strongly agree” and “agree” versus “disagree” and “strongly disagree”) were combined to provide a single percentage representing whether respondents agreed or disagreed with a statement.

## RESULTS

### Participant Characteristics

We sent 2,024 email invitations. In total, 490 individuals accessed the survey. Nine people indicated they did not want to participate. Of the 481 people who indicated they agreed to participate, 74 did not provide answers to any questions, leaving 407 respondents (85%) for analysis and a final response rate of 20% of those invited. Researchers from 39 countries answered the survey. Participant demographics are reported in **Table 1**. Approximately half (54%) of researchers were female, 28% held an MD, and 56% held a PhD without an MD degree. Sixty-six percent indicated that their research roles included “overall study design” and 81% were involved in analysis of genomic samples/data. The vast majority (86%) reported their research involved array-based testing (e.g., SNP arrays), but a substantial number were also using genome sequencing (48%), exome sequencing (38%), and single-gene testing (32%). Participant roles, type of genetic testing used, disorders examined, and patient populations are shown in **Table 2**.

**Table 1.** Psychiatric genetics researchers’ demographics

|  | **% (n)** |
| --- | --- |
| **Total** | 100% (407) |
| **Gender (n=350)** |  |
| Female | 54% (189) |
| Male | 43% (150) |
| I prefer not to say | 3% (11) |
| **Country (n=334)<sup>1</sup>** |  |
| United States | 42% (139) |
| United Kingdom | 10% (32) |
| Canada | 9% (30) |
| Germany | 4% (14) |
| Brazil | 4% (13) |
| Norway | 4% (13) |
| Australia | 3% (11) |
| Sweden | 3% (11) |
| Other European Countries | 15% (51) |
| Asian Countries | 5% (18) |
| Other Countries in the Americas | 4% (12) |
| African Countries | 2% (7) |
| Other Oceania Countries | .3% (1) |
| **Academic degree (n=351)<sup>1</sup>** |  |
| Ph.D. only | 56% (202) |
| M.D. only | 15% (51) |
| M.D. and Ph.D. | 13% (46) |
| M.S., Genetic Counseling only | 3% (10) |
| Other | 12% (42) |
| **Years in psychiatric genetics research (n=343)** |  |
| 0-4 years | 25% (85) |
| 5-9 years | 33% (114) |
| 10-19 years | 26% (88) |
| 20-29 years | 11% (37) |
| 30+ years | 5% (19) |

<sup>1</sup>Respondents could select all responses that applied.

**Table 2.** Psychiatric genetics researchers’ roles, testing, and populations

|  | % (n) |
| --- | --- |
| **Role (n=407)<sup>1</sup>** |  |
| Analysis of genomic samples/data | 82% (332) |
| Overall study design | 66% (270) |
| Collection of clinical data and biospecimens | 38% (154) |
| Generating genomic data | 34% (138) |
| Obtaining informed consent | 26% (104) |
| Providing clinical care | 18% (74) |
| Other | 4% (18) |
| **Genetic Test Used in Research (n=405)<sup>1</sup>** |  |
| Array-based testing (e.g., SNP arrays) | 86% (348) |
| Genome sequencing | 48% (195) |
| Exome sequencing | 37% (152) |
| Single-gene testing | 32% (131) |
| Panel-based testing | 14% (55) |
| Karyotyping | 6% (26) |
| Other | 6% (24) |
| **Psychiatric Disorder Studied (n=352)<sup>1</sup>** |  |
| Schizophrenia and Related Disorders | 55% (193) |
| Depressive Disorders | 41% (146) |
| Bipolar Disorder | 38% (135) |
| Autism Spectrum Disorder | 25% (89) |
| Attention-Deficit Hyperactivity Disorder | 19% (67) |
| Anxiety Disorders | 16% (57) |
| Obsessive-Compulsive Disorder and Related | 12% (42) |
| Alzheimer's Disease | 11% (38) |
| Eating Disorders | 10% (36) |
| Substance Abuse / Addiction | 9% (31) |
| Tourette's Syndrome | 6% (20) |
| Suicide | 2% (7) |
| Huntington's Disease | 2% (6) |
| Other | 9% (32) |
| **Patient Population (n=405)<sup>1</sup>** |  |
| Adults | 94% (382) |
| Children | 44% (179) |
| Adults lacking decision-making capacity | 14% (58) |
| Children not expected to have decision-making capacity as adults | 12% (49) |

<sup>1</sup>Respondents could select all responses that applied.

### Researchers’ Current Return of Results Practices

Most respondents (61%) reported that their studies generate medically relevant psychiatric genomic findings and almost half (46%) generate medically relevant non-psychiatric genomic findings. A quarter of researchers (24%) have worked on studies that have returned individual genomic results to participants (see **Table 3** for frequencies of types of results returned). Of those who have returned results (n=97), 60% have returned medically actionable results related to psychiatric disorders, 38% have returned non-medically actionable results related to psychiatric disorders, and 28% have returned variants of unknown significance (VUS) in genomic loci associated with psychiatric disorders. Notably, individual findings were not always confirmed by a clinical laboratory before being returned to participants, with 26% indicating that findings were not corroborated when returned, and 16% indicating findings were corroborated only sometimes. Half of researchers (52%) involved in decisions about return of results reported that they plan to return or continue to return individual findings.

**Table 3.**
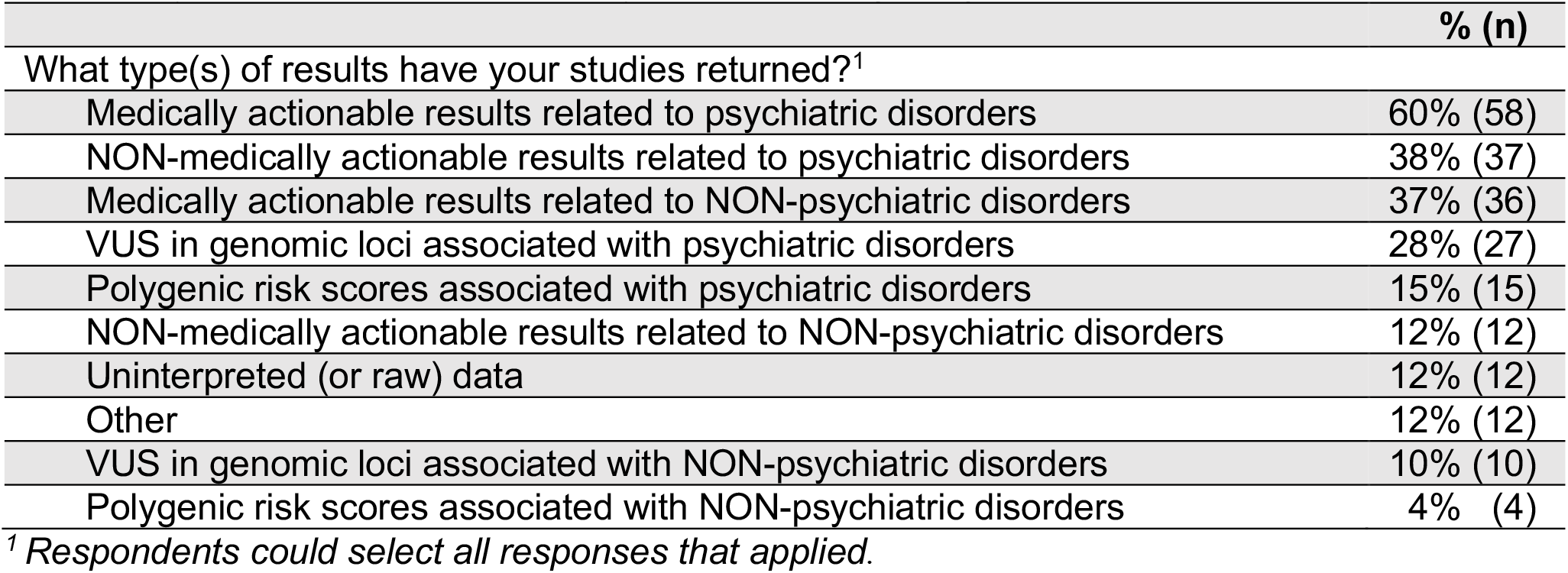
Types of results returned by researchers (n=97)

### Researchers’ Motivations for Returning Research Results to Participants

Researchers were asked about potential motivations for offering individual genomic research results related to psychiatric disorders to patient-participants. The vast majority (86%) agreed that it is the ethical thing to do when the results are medically actionable, while 45% agreed it is the ethical thing to do when the results are medically relevant, even if not actionable. Seventy-six percent agreed that results related to psychiatric disorders should be offered because it can show respect for a patient-participant’s autonomy if the participant wants the results and 73% agreed it recognizes that participants should have ownership over their data. A majority of respondents (73%) agreed results should be offered because it can help patient-participants better understand the role of genetics in psychiatric disorders and 50% agreed that it can reduce stigma around psychiatric disorders. Half (53%) of respondents agreed it can incentivize study participation.

### What Should be Offered?

Researchers were asked their perspectives on which types of findings should be offered to patient-participants (**Figure 1**). Most researchers agreed that medically actionable (see definition above) genomic findings related to psychiatric disorders (82%) and non-psychiatric disorders (81%) should be offered. Fewer researchers agreed that non-medically actionable findings related to psychiatric disorders (32%) and non-psychiatric disorders (32%) should be offered, though more than a quarter of respondents were ambivalent or unsure about both. When asked about specific findings that are typically considered non-medically actionable, however, 71% agreed that results related to risk for Huntington’s disease should be offered, and 64% agreed that results related to risk for Alzheimer’s disease should be offered. Consistently, most researchers (61%) agreed that findings that could be “personally useful (e.g., for planning finances, long-term care, etc.), even if not medically actionable,” should be offered, and 57% agreed that genomic research results related to psychiatric disorders should be offered to patient-participants because it can allow for family and life planning.

**Figure 1.**
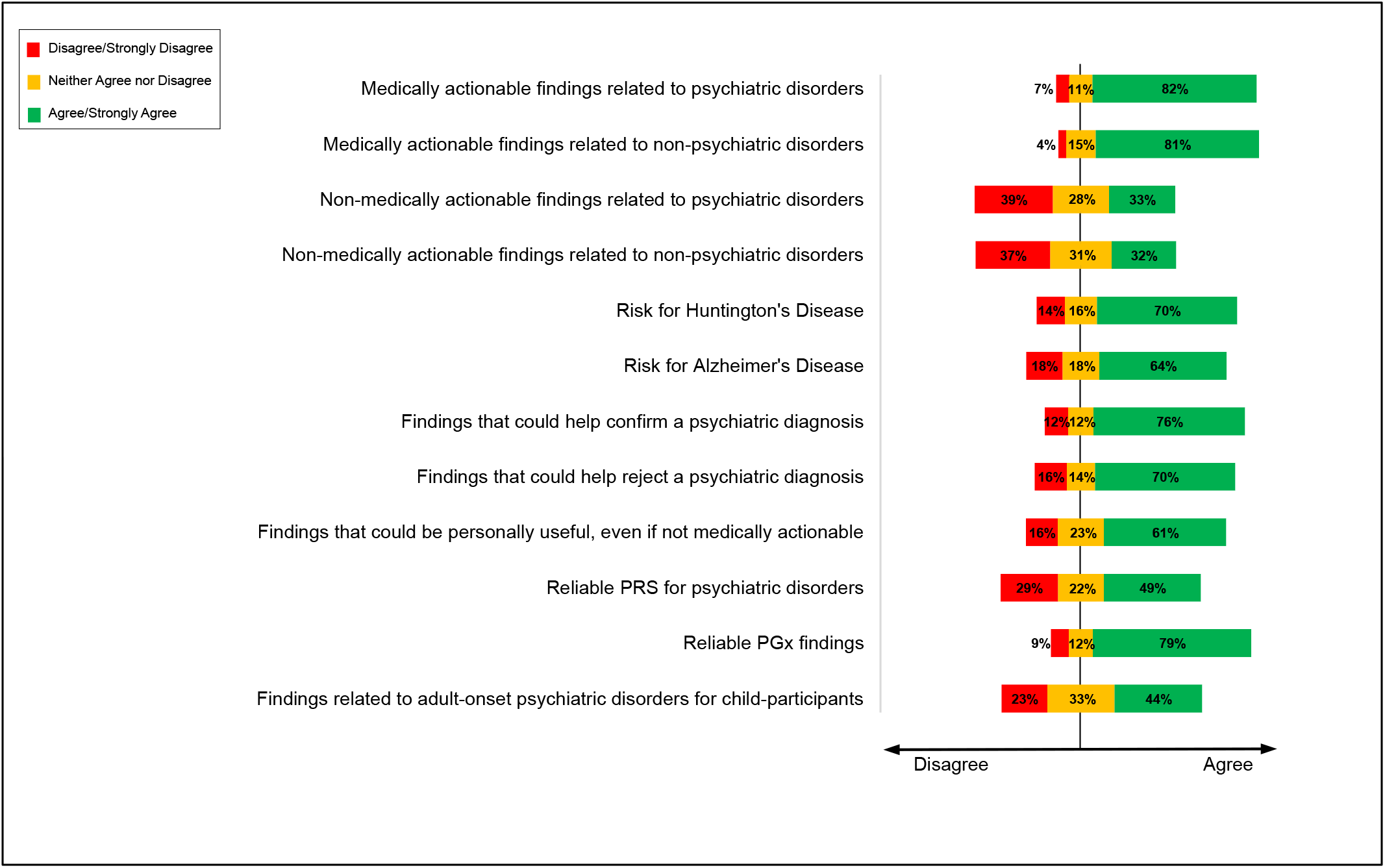
Psychiatric genetics researchers supported offering a variety of potential findings to participants. Each type of result was asked on a 5-point Likert scale anchored by “Strongly Disagree” to “Strongly Agree” with “Neither Agree nor Disagree” as the neutral midpoint.

About half of researchers disagreed that VUS should be offered to patient-participants, even if VUS are identified in genomic loci associated with a participant’s psychiatric symptoms (53% disagree; 26% agree), or a participant’s non-psychiatric symptoms (56% disagree; 21% agree). Similarly, respondents disagreed when asked if VUS in genomic loci associated with a psychiatric (54% disagree; 22% agree) or non-psychiatric (57% disagree; 19% agree) condition should be returned when the patient-participant has no symptoms of the condition. The range of respondents selecting “neither agree nor disagree” was between 22% and 25% in these four VUS items. Most researchers (63%) disagreed with offering uninterpreted (unprocessed or “raw”) data to patient-participants, although 18% supported this.

Researchers were also asked about offering polygenic risk scores (PRS) and pharmacogenetic findings if these were reliable for all populations. Almost half of researchers agreed that reliable PRS for psychiatric disorders (49% agree; 29% disagree) and non-psychiatric disorders (49% agree; 27% disagree) should be offered. Meanwhile, fewer (23%) agreed that PRS related to educational attainment should be offered, and 52% disagreed. The majority of researchers (79%) agreed that reliable pharmacogenetics findings should be offered.

When asked about returning results when children are participants in psychiatric genomic research, 56% of researchers agreed that results related to childhood-onset psychiatric disorders should be offered, 44% agreed that results related to adult-onset psychiatric disorders should be offered, and 37% agreed that reliable PRS related to psychiatric disorders should be offered. Nearly one-third of respondents responded neither agree nor disagree to these questions about offering results when children are participants.

### Knowledge about Return of Results

Despite general support throughout the survey for returning genomic research results to participants, respondents reported a lack of knowledge about how to manage this process responsibly. Most researchers (59%) rated their knowledge about how to manage the entire process for offering and returning results to participants as poor or very poor. When asked about specific aspects of the return of results process, almost half of researchers rated their knowledge as poor or very poor in different areas: selecting which findings to offer (41%); conducting the informed consent process for return of results (43%); selecting how to disclose results (e.g., in person, via telephone; 48%); selecting who will disclose results (41%); and determining what information should be provided to the participant along with the results (48%). Respondents also reported lack of relevant guidelines, with 14% agreeing that there are adequate guidelines for returning genomic research results, and even fewer (9%) indicating that there are adequate guidelines for returning genomic research results specifically related to psychiatric disorders.

## DISCUSSION

### Practices

This is the first survey to examine practices and perspectives toward return of research results in a global sample of psychiatric genetics researchers. Most respondents were working on studies that generated medically relevant findings related to psychiatric conditions, and almost half were generating medically relevant findings related to non-psychiatric conditions. Yet, only a quarter of researchers have returned findings to individual participants. Thus, a significant number of medically relevant findings are being generated, but not currently offered to participants who could potentially benefit from this information. On the other hand, over half of researchers involved in decisions about whether to return findings reported that they plan to offer individual research findings in the future. This suggests that return of results to individual participants is a growing practice in psychiatric genetics research.

Notably, almost half of researchers who indicated they have worked in studies that have returned individual results reported that results were not, or only sometimes, confirmed in a clinical laboratory. This included researchers from the United States, where there has been debate about whether the law that regulates clinical laboratories (Clinical Laboratory Improvements Act; CLIA) allows this.^26^ In fact, the most widely held interpretation of the law is that CLIA does not allow individual return of results by laboratories that are not CLIA certified, ^24,26,27^ and the vast majority of genetics research laboratories in the US are not. These findings suggest that when studying or developing policies about return of results, it should not be assumed that research findings are first being confirmed by clinical laboratories before being returned to participants. Thus, it is important to develop minimum requirements for handling and processing samples, analysis, results, and disclosure.^16,26^ This should include a uniform and clear way of informing participants that they should not make decisions based on research findings that have not been corroborated by a clinical laboratory, and should consult a clinician, ideally a genetic specialist (e.g., genetic counselor, medical geneticist), to evaluate if any next steps are appropriate.

### What Findings Should be Offered

For years, much of the impetus for returning results in genetics research has been that some findings are medically actionable, thus, knowing about these could benefit the participant because there are medical interventions that could decrease the risk of poor health outcomes.^28,29,30^ Consistently, the majority of psychiatric genetics researchers agreed that medically actionable psychiatric and non-psychiatric findings should be offered. They were less supportive of returning non-medically actionable results, yet most agreed that results related to risk for Alzheimer’s and Huntington’s disease, conditions typically considered non-medically actionable, should be offered to patient-participants. Traditionally, there has been some resistance about offering non-medically actionable findings in genetics research. One of the strongest arguments against returning or offering non-medically actionable findings is that the harms may outweigh the benefits, as the information could cause distress but there is no medical intervention that can help minimize the likelihood of poor health outcomes. The respondents’ support for offering these findings suggests a potential shift in researcher attitudes^24^ toward offering this type finding. On the other hand, psychiatric genetics researchers had not been surveyed about this before, thus, it may be that their experience and familiarity with these conditions makes them more comfortable with offering this information to participants.

Psychiatry researchers’ views about offering findings related to risk for Alzheimer’s and Huntington’s, in addition to PRS for psychiatric disorders, may also be explained by having a broader notion of what “actionability” entails. In our previous research with psychiatric genetics researchers, we found evidence for at least four ways in which these researchers use the term actionability.^10,17^ First, researchers use the term in the typical fashion to refer to genomic findings for which there is some type of medical intervention that can help decrease the risk for poor health outcomes. Second, they use the term to refer to how findings may have personal utility in that they can help participants plan finances, housing, and other aspects of life considering their genomic risks. Third, these researchers use actionability to refer to behaviors or lifestyle practices such as good sleep hygiene or exercise that could potentially help decrease risk, severity, and future episodes^31^ of psychiatric disorders. Finally, psychiatric genetics researchers also use the term actionability to refer to the possibility of using medical interventions to minimize poor health outcomes from other phenotypes associated with a genomic variant known to also increase the risk for psychiatric disorders (e.g., calcium supplementation in patients with 22q11.2 deletion syndrome).^32^

Recently, there has been a significant amount of debate about the utility and ethical implications of using PRS in psychiatry.^33,34,35,36^ While some see PRS as a potential tool to help prevent and manage care for psychiatric disorders at both an individual and general population level, others question its utility at an individual level.^35^ Respondents in the present study are likely some of the most knowledgeable experts about the utility and limitations of PRS in psychiatry. Some of them reported that they are returning PRS for psychiatric disorders and about half agreed that if PRS were generated in the course of research and these were reliable for all populations, they should be offered to participants. This suggest that researchers see some value for the use of psychiatric PRS at an individual level. More research is necessary to explore the perspectives of expert stakeholders, clinicians, patients, and research participants regarding the utility and ethics of using PRS for psychiatric disorders in different settings.

Interestingly, the majority of respondents were either supportive or ambivalent about offering research results related to adult-onset psychiatric disorders when children participate in psychiatric genomic research. While professional society guidelines support clinical genetic testing and genome-scale sequencing for children for specific reasons, disclosing predictive genetic results for late-onset conditions has historically been discouraged, stemming from appeals to the child’s “right to an open future” and concerns about the psychosocial impact of this information.^37,38^ Our results show that psychiatric genetics researchers are largely not opposed to returning such information when the results are related to psychiatric disorders. This may be due to their broader perspective of what actionability entails and because the average age of onset for many psychiatric conditions is late adolescence and early adulthood, and early surveillance and intervention could potentially improve clinical outcomes.^39,40^ Future research should explore this finding to better understand psychiatric genetics researchers’ perspectives toward the risks and benefits of disclosing such information to children and their families.

### Opportunities Moving Forward

Most respondents did not believe there are adequate guidelines for returning genomic research results related to psychiatric disorders and reported having poor knowledge about how to manage the process of returning results. These results suggest that there is an opportunity for professional organizations to develop guidelines for determining whether and which results should be offered and how the information should be delivered. In addition, it would be helpful to create decision aids to help researchers explain to participants the range of results that may be generated and help participants decide whether they want this information.

### Limitations

The study sample comprised a diverse group of researchers across 39 countries, but it is possible that these results may not be representative of the larger population of psychiatric genetics researchers. Survey respondents self-selected for participation in this study and it is possible that researchers who were more positive toward or more knowledgeable about return of research results may have been more likely to participate. Additionally, as there is a growing consensus toward return of research results to participants, there is the potential for social desirability bias with some of our questions, including those asking about current practice of return and attitudes toward returning results. Despite these limitations, this is the first study to assess practices of and perspectives toward return of research results in a global sample of psychiatric genetics researchers and thus contributes much needed empirical evidence that can help shape future practice and guidelines around the return of research results in psychiatric genetics research.

## CONCLUSIONS

Return of results is a growing practice in psychiatric genetics research. As research in psychiatric genetics continues to grow and increasingly utilize more comprehensive SNP arrays and genome sequencing technologies, the potential for researchers to generate medically relevant findings that researchers want or feel compelled to return will increase. Our findings suggest that psychiatric genetics researchers may be supportive of returning a wide range of genomic research results to participants, but that they feel they lack adequate knowledge and guidance to do so responsibly. This deficiency represents an opportunity for relevant professional organizations to provide guidance and for future research to assess best practices for returning results to this patient population responsibly.

## Data Availability

The datasets generated during and/or analyzed during the current study are available from the corresponding author on reasonable request.

## Acknowledgments

Research for this article was funded by the National Human Genome Research Institute (NHGRI) of the National Institutes of Health (NIH) Grant R00HG008689 (Lázaro-Muñoz, G). The views expressed are those of the authors alone, and do not necessarily reflect views of NIH or Baylor College of Medicine.

